# Amyloid and tau pathologies are drivers of white matter damage in aging and Alzheimer’s disease

**DOI:** 10.64898/2025.12.18.25342588

**Authors:** Farooq Kamal, Mahsa Dadar

**Affiliations:** Department of Psychiatry, McGill University, 1033 Pine Ave W, Montreal, Quebec, H3A 1A1, Canada; Douglas Mental Health University Institute, 6875 Bd LaSalle, Verdun, Quebec, H4H 1R3, Canada

**Keywords:** mild cognitive impairment, amyloid, tau, White Matter Hyperintensities, AD

## Abstract

**BACKGROUND:** White matter hyperintensities (WMHs) are increasingly recognized as markers of cerebrovascular pathology in Alzheimer’s disease (AD), yet their temporal relationship with amyloid and tau accumulation remains unclear. While previous studies suggest bidirectional associations between WMHs and AD pathology, regional associations between WMHs and AD pathology have yet to be examined. This study investigated the temporal and regional associations between PET measures of amyloid (Aβ) and tau pathology and WMH burden in older adults.

**METHODS:** Data from the Alzheimer’s Disease Neuroimaging Initiative (ADNI) included 1,241 older adults with Aβ and 636 with tau for cross-sectional analyses. Longitudinal analyses included 670 participants for Aβ change and 1,079 for WMH change (Aβ group), and 199 for tau change and 356 for WMH change (tau cohort). Linear models were used to i) assess associations between baseline regional WMH and Aβ and tau pathology, and ii) examine whether baseline pathology in one measure was associated with change in the other measure over two years.

**RESULTS:** Baseline analyses revealed significant bidirectional associations between WMH burden and both Aβ (*t*=2.09-4.16, *p*<.05) and tau pathology (*t*=2.44-2.87, *p*<.04), with stronger effects in posterior brain regions. Longitudinal analyses showed that baseline Aβ levels were associated with future WMH progression in frontal and occipital regions (*t*=2.44-3.27, *p*<.03), while baseline tau was linked to WMH increases in frontal and parietal regions (*t*=2.48-3.51, *p*<.03). However, baseline WMH burden was not associated with future accumulation of either Aβ or tau pathology in any region.

**CONCLUSIONS:** These findings suggest that Aβ and tau pathology drive future WMH progression rather than the reverse, with distinct regional patterns for each pathology type.

## 1. Introduction

Alzheimer’s disease (AD) is typically characterized by amyloid-βeta (Aβ) plaques and neurofibrillary tau tangle buildup in the brain.^1–5^ Although Aβ and tau are hallmarks of AD, cerebrovascular pathology, including white matter hyperintensities (WMHs) is also a highly prevalent pathology associated with the disease.^6–9^ WMHs which appear as bright spots on T2-weighted and fluid-attenuated inversion recovery (FLAIR) magnetic resonance imaging (MRI), reflecting small-vessel disease (SVD) and are characterized by demyelination and axonal loss.^8–11^ These lesions have been associated with increased risks of cognitive decline across the spectrum from healthy aging to AD. ^6,12,13^

What remains less clear is whether cerebrovascular pathology occurs before, after, or concurrently with the progression of Aβ and tau pathology, and whether these temporal relationships differ across brain regions. Recent longitudinal studies have suggested that cerebrovascular pathology might initiate a cascade of events leading to AD neurodegeneration and cognitive decline.^8,9,14,15^ On the other hand, Aβ deposition may increase WMH burden through processes that are not necessarily vascular in nature, including neuroinflammation and oxidative stress.^16,17^ Under this model, an initial rise in Aβ would damage white matter, which would further elevate Aβ levels, creating a cyclical process of mutual reinforcement.^18–20^ Similarly, tau pathology may contribute to white matter damage through direct toxic effects or secondary inflammatory processes.^21,22^

Previous work by our group^8^ provided insights into the temporal relationships between WMHs, neurodegeneration, and cerebrospinal fluid (CSF) Aβ_1-42_ levels, suggesting that baseline WMHs are associated with future gray matter atrophy and cognitive decline, while baseline Aβ_1-42_ levels are also associated with future WMH increase. However, this analysis was limited to whole-brain WMH and gray matter atrophy measures and CSF Aβ biomarkers and did not examine tau, leaving important questions about regional variations and direct brain pathology measurements unanswered. It is well-established that both AD pathology and WMH show non-uniform spatial distributions throughout the brain.^14,15,23^ Aβ deposition typically begins in neocortical regions, while tau pathology progresses through specific neurofibrillary stages, starting from the medial temporal lobes. ^24–27^ In addition, WMHs often show a posterior predominance in AD associated with greater cognitive impact.^14–16,28^

The present study aimed to extend our previous findings^8^ by examining regional temporal relationships between PET-measured Aβ and tau pathology and WMH burden using longitudinal data. Specifically, we investigated: (1) whether baseline WMH burden is associated with future accumulation of Aβ and tau pathology in different brain regions; (2) whether baseline regional Aβ and tau pathology levels are linked to future WMH accumulation; and (3) whether these temporal relationships differ between anterior and posterior brain regions. By addressing these questions, we sought to clarify the sequence of pathological events in AD and identify potential regional differences that could inform our understanding of disease progression.

## 2. Methods

### 2.1 Alzheimer’s Disease Neuroimaging Initiative

The data used in this study were obtained from the Alzheimer’s Disease Neuroimaging Initiative (ADNI) database (adni.loni.usc.edu). ADNI was launched in 2003 as a public-private partnership, led by Principal Investigator Michael W. Weiner, MD. The dataset’s primary objective is to determine whether serial MRI, positron emission tomography (PET), other biological markers, and clinical and neuropsychological assessments can be combined to measure the progression of mild cognitive impairment (MCI) and early Alzheimer’s disease (AD). The study was approved by the institutional review boards of all participating institutions, and written informed consent was obtained from all participants or their authorized representatives. Participants were selected from ADNI-1, ADNI-2, ADNI-GO, and ADNI-3 cohorts based on availability of neuroimaging data and clinical assessments. Detailed inclusion and exclusion criteria are available at www.adni-info.org.

### 2.2 Participants

All participants were aged 55-90 years at the time of recruitment, who had available apolipoprotein E ε4 (APOE4) genotype information, and baseline and follow-up MRI (from which WMHs can be extracted) and PET (tau and Aβ) data. At baseline, 1,241 older adults with Aβ data and WMH data, and 636 older adults with tau data and WMH data were included. These baseline analyses examined the cross-sectional associations between WMH burden and PET (Aβ and tau).

For longitudinal analyses, participants were included if they had baseline measures and follow-up data at approximately 2 years post-baseline. Two separate longitudinal subsamples were established based on the variable data. When examining change in PET (Aβ and tau) as the outcome (baseline WMH associations with future PET changes), 670 participants with Aβ data and 199 participants with tau data were included. When examining change in WMH as the outcome (baseline PET associations with future WMH changes), 1,079 participants with baseline Aβ data and 356 participants with baseline tau data were included. The difference in longitudinal sample sizes reflects the availability of follow-up data for each outcome.

### 2.3 PET Imaging Acquisition and Processing

Regional Aβ PET (florbetapir) standardized update value ratios (SUVRs) and regional tau PET (flortaucipir) SUVRs were downloaded from ADNI. Aβ PET imaging was performed using [^18^F]-florbetapir (AV-45), while tau PET utilized [^18^F]-flortaucipir (FTP). Standardized ADNI protocols were employed for all PET acquisitions, with detailed parameters available at adni.loni.usc.edu. Additionally, the ADNI pipeline was utilized to preprocess the Aβ and tau PET imaging data, including coregistering the PET scans to the structural MRI, and FreeSurfer was used to generate regional data.^29–31^ Regional SUVRs were calculated using volume-weighted averaging of bilateral cortical parcellations derived from FreeSurfer processing. Brain regions were defined as follows: frontal (including prefrontal, orbitofrontal, precentral, and paracentral regions), temporal (encompassing entorhinal, fusiform, inferior/middle/superior temporal, and parahippocampal areas), parietal (comprising inferior/superior parietal, supramarginal, postcentral, precuneus, and posterior cingulate regions), and occipital (lateral occipital, cuneus, lingual, and pericalcarine areas). Anterior and posterior composite measures were derived by averaging frontal and temporal lobe SUVR (anterior) and parietal and occipital lobe SUVR (posterior) regions, respectively.

### 2.4 Structural MRI Acquisition and WMH Measurements

Structural T1-weighted MRI scans were acquired using standardized ADNI protocols. MRI protocols and imaging parameters can be found at: http://adni.loni.usc.edu/methods/mri-tool/mri-analysis/. All participant MRI data (baseline and longitudinal) were downloaded from the ADNI public website. All T1w scans were pre-processed using our standard^32^ which includes: noise reduction, ^33^ intensity inhomogeneity correction, ^34^ and intensity normalization into range [0-100]. These pre-processed images were then linearly (9 parameters: 3 translation, 3 rotation, and 3 scaling) ^35^ registered to the MNI-ICBM152-2009c average template. ^36^

### 2.5 WMH measurements

WMH segmentation was performed based on T1-weighted images using a validated automated technique previously tested in aging and neurodegenerative populations to obtain WMH measurements. ^37^ The technique has been validated in multi-center studies ^38,39^ and within the ADNI cohort. ^40^ T1-weighted images were used instead of FLAIR or T2-weighted scans since ADNI1 only collected T2-weighted images at 1×1×3 mm^3^ resolution, ADNI2 and ADNI-GO only had axial FLAIR images at 1×1×5 mm^3^ resolution, and ADNI3 acquired sagittal FLAIR images at 1×1×1.2 mm^3^ resolution. These different resolutions would make WMH comparisons unreliable across ADNI sub-studies. Thus, we opted to use T1-weighted images that were consistently acquired to estimate WMH burden. Furthermore, we have previously shown that T1w-based WMH volumes correlate strongly with FLAIR and T2w-based measurements in ADNI dataset.^41^ The quality of the registrations and WMH segmentations was visually assessed (by M.D.), and cases that failed quality control were removed from the analyses. WMH load was defined as the volume of all voxels identified as WMH in the standard space (in mm^3^) and were thus normalized for head size. Both regional and total WMH volumes were calculated using the Hammers Atlas. ^37,41^ All WMH volumes were log-transformed to achieve normal distribution. Regional WMH values (i.e., frontal, temporal, parietal, and occipital) were averaged across the right and left hemispheres to obtain one measure for each region. Given that WMHs often show a posterior predominance in AD pathology and follow a posterior to anterior shift, we created anterior composites (averaged using frontal and temporal regions) and posterior composites (averaged using parietal and occipital regions) for both WMHs and PET measures (tau and Aβ).

### 2.6 Statistical Analysis

Demographic group differences were assessed using independent sample *t*-tests for continuous variables including age, education, and chi-square (*x*^2^) tests for categorical measures including sex and APOE4 carrier status. The categorical variable APOE4 was used to contrast participants with one or two APOE ɛ4 alleles against those with zero.

Baseline PET scans were paired with the closest MRI scan within ±365 days. All models were examined for each of the four lobes (frontal, temporal, parietal, occipital) including anterior (frontal+temporal) and posterior (parietal+occipital) composites. Baseline and longitudinal models included age at baseline, education, sex, diagnostic group status, and APOE4 status as covariates.

#### 2.6.1 Baseline Cross-sectional Analyses

For baseline analyses, models were run for each regional PET (tau and Aβ) measure and WMH separately. First, linear regression models examined associations between baseline WMH burden and PET levels in matching regions, with PET SUVRs as outcomes.

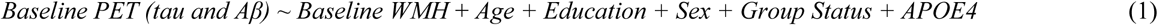

#### 2.6.2 Baseline Cross-sectional Analyses in Common Subset

To verify that baseline findings were not driven by differences in cohort composition between Aβ and tau groups, baseline analyses were repeated in a common subset of participants with complete baseline data for Aβ, tau, and WMH. Baseline cross-sectional models (Equation 1) for both Aβ and tau were repeated in this common cohort to assess whether the regional patterns of associations remained consistent with the main analyses.

#### 2.6.2 Longitudinal Analyses

Longitudinal models examined the temporal relationships between PET (Aβ and tau) and WMH burden. Longitudinal change was calculated using follow-up scans closest to 2 years post-baseline. To ensure temporal consistency and minimize the influence of differences in follow-up durations, we restricted our longitudinal sample to participants with follow-up intervals between 1 and 3 years (inclusive). To further account for variability in follow-up intervals, change in each measure was annualized by dividing the total change by the follow-up interval in years (Δ measure = [value at follow-up – value at baseline] / interval in years). This annualized change was then modeled as a function of baseline values while controlling for covariates. When examining change in PET (Aβ or tau) as outcomes:

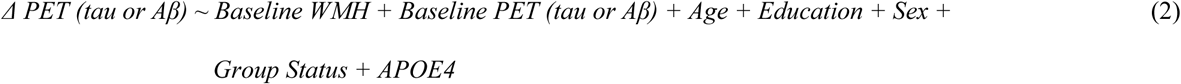

To examine whether baseline PET Aβ or tau were associated with future WMH accumulation, we used:

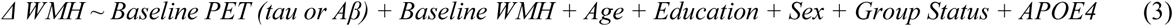

Aβ and tau analyses were conducted separately due to limited overlap in participants with both PET modalities at baseline and follow-up timepoints.

#### 2.6.3 Anterior-Posterior longitudinal Analysis

To test whether the associations between tau and Aβ and WMH differed across anterior and posterior regions, we contrasted their slopes using an interaction term in the linear models (4). Specifically, we tested whether the relationship between the primary baseline measure (baseline WMH or baseline PET) and the longitudinal outcome (Δ PET or Δ WMH) varied by region (Anterior vs Posterior), using an interaction term between the baseline measure and a region-grouping factor. Thus, to test whether baseline WMH was associated with longitudinal change in PET across anterior and posterior regions, the following model was used:

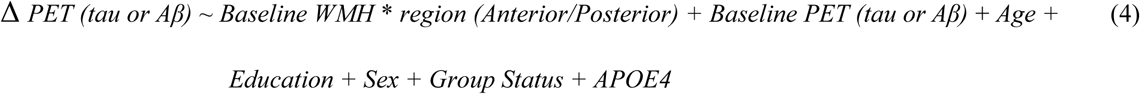

To test whether baseline PET was associated with longitudinal change in WMH across anterior and posterior regions, the following model was used:

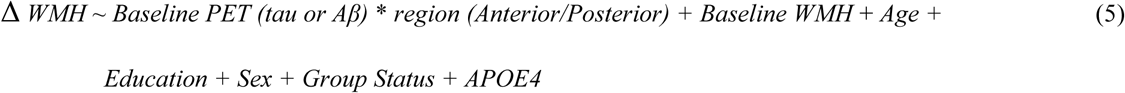

Region was treated as a categorical variable (Anterior vs Posterior), and the interaction term tested whether the strength of association differed by region. All analyses were carried out in R. To normalize the data for analysis, all SUVR, WMH and continuous values were log-transformed and z-scored. Multiple comparison correction was applied using false discovery rate (FDR) procedures. ^42^ All p-values reported are the FDR corrected p-values.

## 3. Results

Demographic and clinical characteristics of the participants are included in Table 1. Participants were grouped based on the availability of Aβ or tau PET data for each analysis. In contrast to baseline, longitudinal demographics did not match between the ΔPET group and ΔWMH group.

**Table 1:**
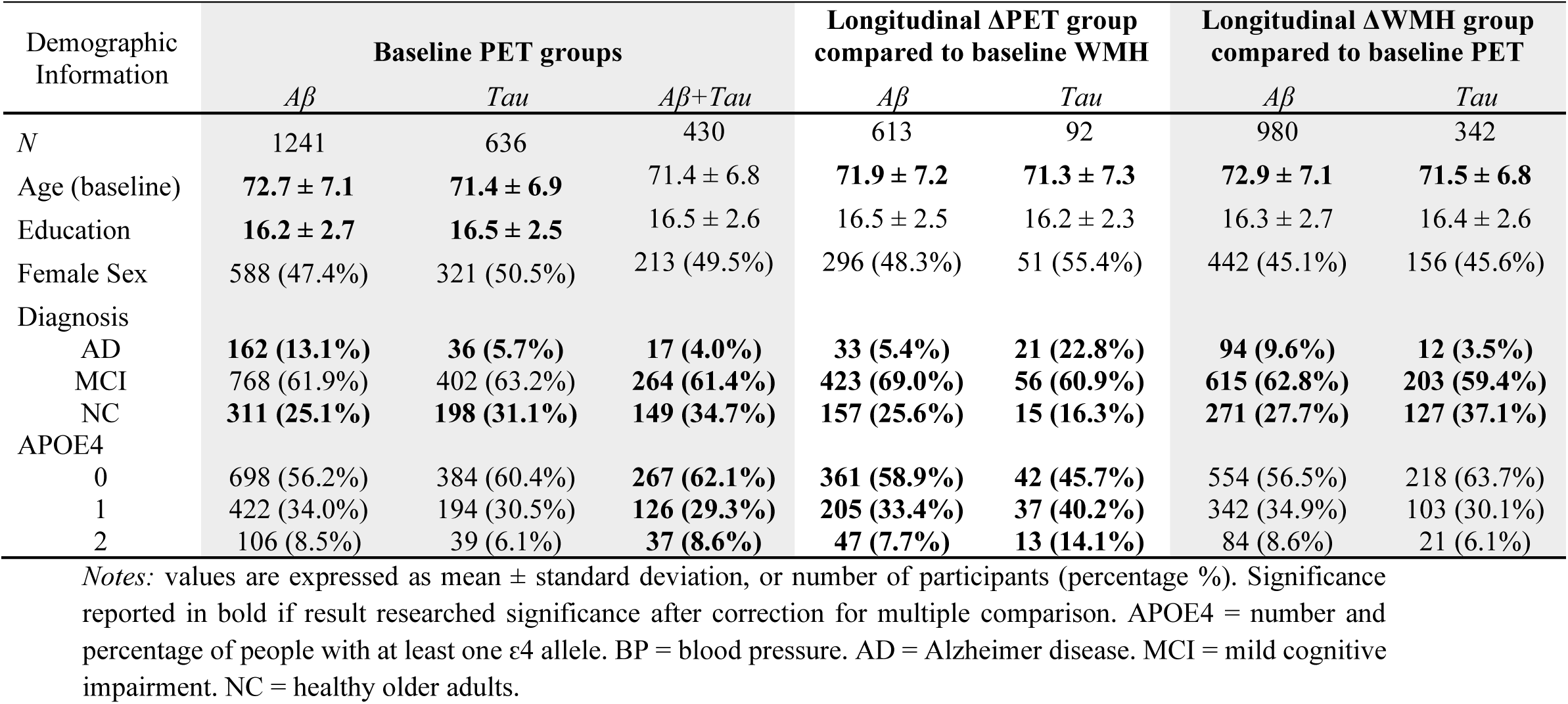
Demographic and clinical characteristics for PET Aβ and Tau for baseline and longitudinal

### 3.1 Demographic results

#### 3.1.1 Participants with baseline Aβ vs Tau PET measures

The subset of the participants that had baseline Aβ PET information (*N=*1,241) was older than the subset with baseline tau PET data (*N=*636; *t=*3.95, *p<*.001) and had slightly lower education (*t=*−2.34, *p=*.02). There were no group differences in sex (*p>*.05). Diagnostic group status also differed (χ²=27.65, *p<*.001), with more older adults with AD in the Aβ subset and with fewer cognitively healthy older adults (CN) compared to the tau subset. The proportion of older adults with MCI did not differ between subsets. There were no group differences for APOE4 status (*p*>.05). All included participants in both subsets also had WMH information available.

#### 3.1.2 Participants with longitudinal Aβ vs Tau PET measures

The subset of participants that had longitudinal Aβ PET information (*N*=613) did not differ in age from the subset with longitudinal tau measures available (*N*=92; *t*=0.74, *p*=.46). Education (*t*=1.10, *p*=.27) and sex (*χ*²=1.64, *p*=.20) also did not differ between groups. Diagnostic group status significantly differed (*χ²=*35.4, *p*<.001), with the tau subset showing a substantially higher proportion of older adults with AD, and the proportion of cognitively healthy older adults was lower in the tau subset compared to the Aβ subset. The proportion of older adults with MCI did not differ substantially between groups. APOE4 status also significantly differed between groups (*χ²*=18.2, *p*<.001), with the tau subset showing a lower proportion of APOE4 non-carriers and a higher proportion of APOE4 carriers.

#### 3.1.3 Participants with longitudinal WMH and baseline Aβ vs Tau PET measures

In the subset of participants that had longitudinal WMH information, those with baseline Aβ available (*N*=980) were older than those with baseline tau (*N*=342; *t*=3.18, *p*=.002). Education and sex did not differ between groups (*p>*.05). Diagnostic group status significantly differed between groups (*χ²*=19.7, *p*<.001), with more older adults with AD and fewer cognitively healthy older adults in the Aβ subset. The proportion of older adults with MCI did not differ between groups. There were no group differences for APOE4 status (*p*=.053).

### 3.2 Baseline Analyses

Table 2 summarizes the results of the cross-sectional models. In models with Aβ as the outcome and the same regional WMH as the independent variable, higher WMH burden was significantly associated with higher Aβ levels in temporal (*t =* 2.29, *p =* .02), parietal (*t =* 3.39, *p* < .001), and occipital (*t =* 3.28, *p* < .001) regions. Composite measures showed the same pattern. Higher posterior WMH was associated with higher posterior Aβ (*t =* 3.70, *p <* .001). No significant association was found in the frontal and anterior regions (*p* > 0.05).

**Table 2:**
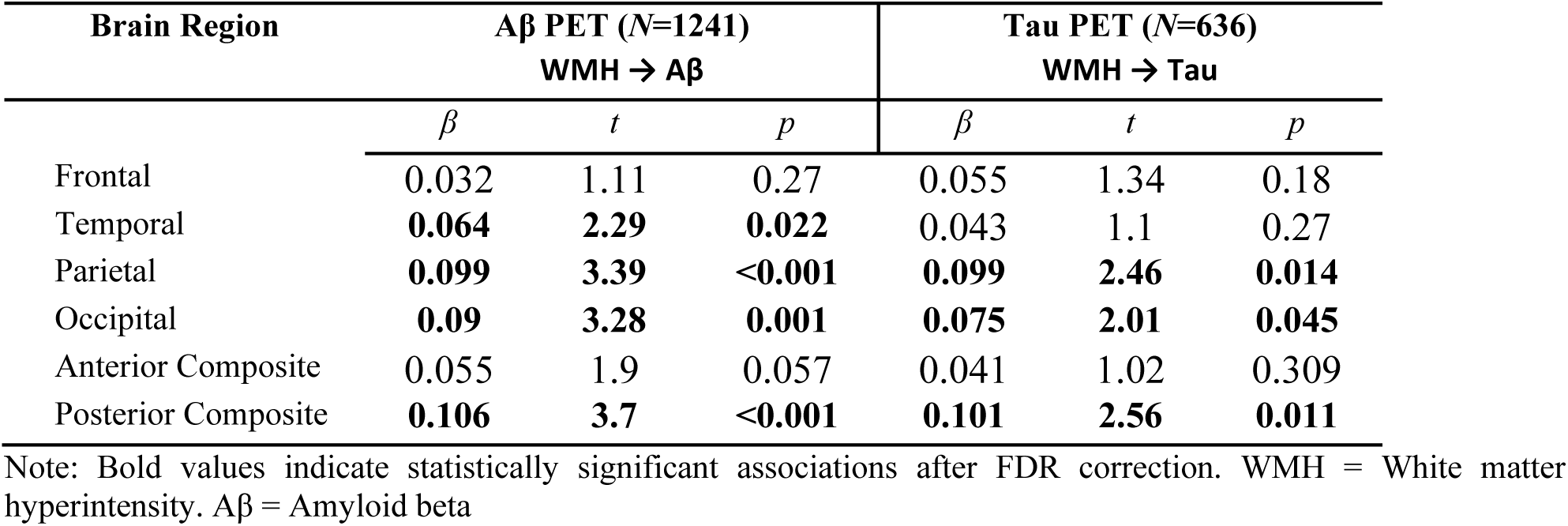
Baseline associations involving WMH with Aβ and Tau across brain regions

In models with tau as the outcome and same regional WMH as the independent variable, higher WMH was associated with higher tau in the parietal (*t =* 2.46, *p =* .014) and occipital (*t =* 2.01, *p =* .045) lobes, and for the posterior composite (*t =* 2.56, *p =* .011). Frontal, temporal, and the anterior composite were not statistically significant (*p* > .05).

To verify that baseline findings were not driven by differences in group composition between Aβ and tau groups, baseline analyses were repeated in a common subset of participants with complete baseline data for Aβ, tau, and WMH (see Table 3). For Aβ, the posterior pattern remained robust in this common cohort. Higher WMH burden was significantly associated with higher Aβ levels in the occipital region (*t =* 3.52, *p* < .001) and the posterior composite (*t =* 2.54, *p =* .011). Associations in other regions (frontal, temporal, parietal, anterior composite) did not reach statistical significance (all *p* > .05), likely due to the relatively smaller sample size of this subset. For tau, the posterior pattern showed a similar trend. Higher WMH was associated with higher tau in the occipital region (*t =* 2.52, *p =* .012) and posterior composite (*t =* 2.37, *p =* .018), although these associations did not survive FDR correction. These findings are consistent with the posterior regional patterns observed in the full baseline analyses.

**Table 3:** Baseline associations involving WMH with Aβ and Tau across brain regions in a common subset

| Brain Region | Aβ PET (N=430) | | | Tau PET (N=430) | | |
| --- | --- | --- | --- | --- | --- | --- |
| | WMH → Aβ | | | WMH → Tau | | |
| | $\beta$ | $t$ | $p$ | $\beta$ | $t$ | $p$ |
| Frontal | 0.011 | 0.23 | 0.82 | 0.068 | 1.37 | 0.172 |
| Temporal | 0.06 | 1.29 | 0.199 | 0.041 | 0.85 | 0.397 |
| Parietal | 0.075 | 1.55 | 0.122 | 0.086 | 1.78 | 0.075 |
| Occipital | **0.16** | **3.52** | **<0.001** | **0.113** | **2.52** | **0.012** |
| Anterior Composite | 0.035 | 0.73 | 0.464 | 0.052 | 1.04 | 0.298 |
| Posterior Composite | **0.121** | **2.54** | **0.011** | **0.111** | **2.37** | **0.018** |

Note: Bold values indicate statistically significant associations after FDR correction. WMH = White matter hyperintensity. Aβ = Amyloid beta

### 3.3 Longitudinal Analyses

Table 4 summarize the results of the longitudinal models. In these models, baseline Aβ levels were significantly associated with greater increase in future WMH burden. With change in WMH as the outcome and baseline Aβ as the independent variable, higher Aβ at baseline was associated with greater WMH burden increase (over 1-3 years) in the frontal (*t =* 3.09, *p =* .002) lobe, as well as in the anterior (*t =* 2.43, *p =* .015) and posterior (*t =* 2.13, *p =* .034) composites. The parietal lobe also showed a significant association (*t =* 2.07, *p =* .039). Temporal and occipital effects did not reach significance. In contrast, when longitudinal change in Aβ was the outcome and baseline WMH the independent variable, baseline WMH burden was not associated with the rate of future Aβ accumulation in any brain region (all *p* > .05).

**Table 4:** Longitudinal associations involving WMH associations with Aβ and Tau across brain regions

| Brain Region | Aβ PET | | | | | | Tau PET | | | | | |
| --- | --- | --- | --- | --- | --- | --- | --- | --- | --- | --- | --- | --- |
| | Baseline WMH $\rightarrow \Delta$Aβ<br>($N = 613$) | | | Baseline Aβ $\rightarrow \Delta$WMH<br>($N=980$) | | | Baseline WMH $\rightarrow \Delta$Tau<br>($N=92$) | | | Baseline Tau $\rightarrow \Delta$WMH<br>($N=342$) | | |
| | $\beta$ | $t$ | $p$ | $\beta$ | $t$ | $p$ | $\beta$ | $t$ | $p$ | $\beta$ | $t$ | $p$ |
| Frontal | 0.008 | 0.17 | 0.865 | **0.105** | **3.09** | **0.002** | -0.017 | -0.14 | 0.886 | 0.108 | 1.92 | 0.056 |
| Temporal | 0.037 | 0.82 | 0.415 | 0.054 | 1.52 | 0.13 | 0.168 | 1.42 | 0.161 | 0.085 | 1.46 | 0.145 |
| Parietal | 0.022 | 0.48 | 0.634 | **0.071** | **2.07** | **0.039** | 0.209 | 1.73 | 0.087 | **0.17** | **2.98** | **0.003** |
| Occipital | 0.04 | 0.96 | 0.338 | 0.05 | 1.49 | 0.136 | 0.027 | 0.24 | 0.807 | -0.044 | -0.76 | 0.447 |
| Anterior Composite | 0.019 | 0.42 | 0.678 | **0.084** | **2.43** | **0.015** | 0.088 | 0.72 | 0.476 | 0.113 | 1.96 | 0.051 |
| Posterior Composite | 0.04 | 0.9 | 0.368 | **0.072** | **2.13** | **0.034** | 0.146 | 1.22 | 0.228 | 0.083 | 1.44 | 0.151 |

Note: Bold values indicate statistically significant associations after FDR correction. WMH = White matter hyperintensity. Aβ = Amyloid beta

A significant temporal relationship was also found for baseline tau pathology associations with subsequent WMH progression. Higher baseline tau levels were significantly associated with greater WMH progression in the parietal region (*t =* 2.98, *p =* .003). The frontal lobe (*t =* 1.92, *p =* .056) and anterior composite (*t =* 1.96, *p =* .051) showed non-significant trends approaching significance. The temporal region, occipital region, and posterior composite showed no significant associations with WMH progression. Similar to the Aβ findings, with change in tau as the outcome and baseline WMH as the independent variable, no significant associations were observed in any region (all *p* > .05). The directionality and regional specificity of these baseline and longitudinal associations are summarized in Figure 1.

**Figure 1.**
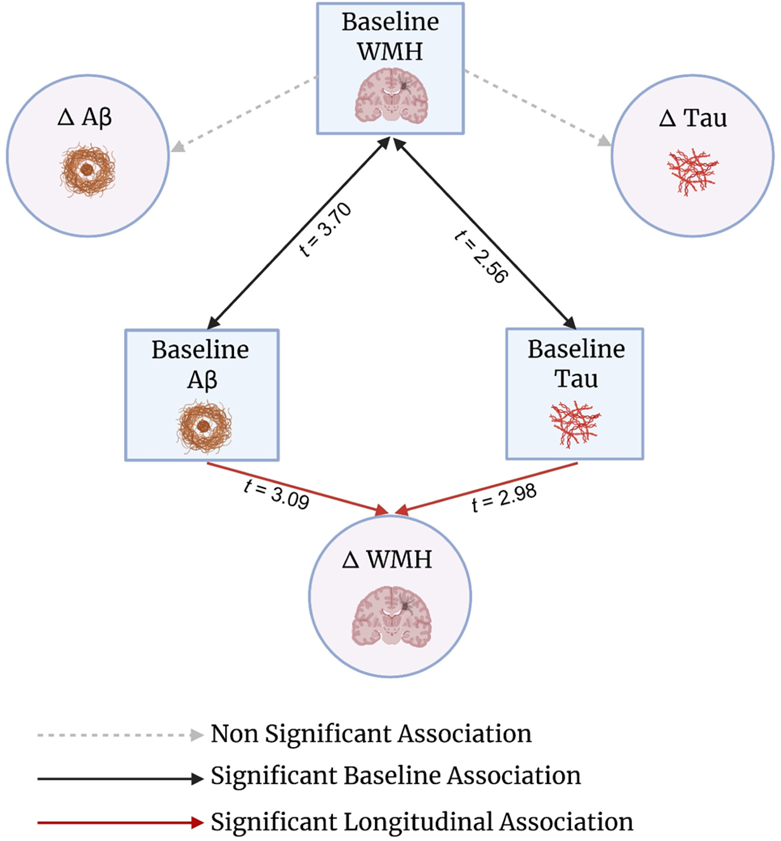
Conceptual model of the directional relationships between WMH and PET pathology. The black arrows summarize cross-sectional findings at baseline: WMH is positively associated with amyloid PET (with strongest effects in posterior regions) and shows significant associations with tau PET (parietal/posterior/occipital). Red arrows summarize the longitudinal directionality: higher baseline amyloid and tau are associated with greater accumulation of WMH over ∼2 years (representative FDR-adjusted p values shown for the strongest effects), whereas the dashed gray arrows indicate that baseline WMH was not associated with subsequent change in amyloid or tau PET.

## Discussion

The present study aimed to examine the regional and temporal relationships between PET-measured Aβ pathology and tau pathology, and WMH burden. While prior work had shown global associations between these measures, research in examining the regional specificity of these associations is limited. Higher WMH burden was found to be significantly associated with increased Aβ in temporal, parietal, and occipital regions, as well as in posterior composite regions. Similarly, tau showed significant associations with WMH in parietal and occipital regions and the posterior composite regions. Additionally, these posterior regional results remained consistent, when analyses were repeated in a common subset of participants with baseline measures for WMH, Aβ, and tau. These findings align with previous studies demonstrating relationships between WMH and AD pathology.^23,43–45^. Additionally, longitudinal analyses demonstrated that baseline Aβ and tau levels were associated with future WMH progression across multiple brain regions. Specifically, baseline Aβ levels were associated with future WMH accumulation in frontal and occipital regions, as well as in anterior and posterior composites. Similarly, baseline tau PET levels showed even stronger associations with future WMH burden, particularly in frontal, parietal, and anterior composite regions. However, baseline WMH burden was not associated with future changes in either Aβ or tau pathology. These results suggest that amyloid and tau pathology precedes or drives white matter changes rather than the reverse, providing new insights into the temporal sequence of pathological events in AD.

These results expand upon our previous findings^8^ based on CSF markers showing that baseline CSF Aβ levels were associated with WMH increase over one year, while baseline WMH was not associated with Aβ changes. Additional studies have further supported the relationship between Aβ and WMH. Walsh et al.^46^ demonstrated that CSF Aβ was consistently associated with WMHs across the disease course from aging to AD, supporting the role of Aβ in exacerbating white matter damage. In contrast, Moscoso et al.^19^ found that baseline WMH burden was associated with longitudinal cortical Aβ accumulation up to 8 years in 190 cognitively healthy, amyloid-negative older adults. However, this relationship was observed at the earliest disease stages and longer follow-up intervals, unlike the present study which included MCI and AD in addition to cognitively healthy older adults and a substantially shorter follow up interval.

The present study extends these previous findings by using regional PET imaging and by including tau pathology, which was not examined in previous studies.^8,46^ Although in these studies, the primary focus was not regional PET or tau pathology, our posterior-dominant associations between WMH and Aβ are consistent with the spatial relationship between amyloid angiopathy and Aβ plaque burden, both of which have previously shown a posterior bias. ^47–49^ The prevalence of protein pathology in the posterior region is consistent with the typical spatial patterns of AD pathology as amyloid plaques often show high accumulation in posterior cortical regions.^50–52^ Our findings also extend previous work reporting that Aβ relates to specific WMH topographic patterns, though they found no relationship between WMH and tau burden. ^53^

Baseline tau was also found to be associated with future WMH burden in the frontal, parietal, and the anterior composite, whereas baseline WMH was not associated with subsequent tau buildup. Previous research has shown mixed findings regarding the relationship between tau pathology and WMH burden.^18,53^ Nonetheless, some neuropathological evidence has shown that cortical tau burden is independently associated with WMH severity in some brain regions, and that the relationship between AD related pathology contributes to parietal and frontal white matter lesions.^54,55^ These results further support models in which cortical tau related neurodegeneration are associated with WMHs.^56,57^ Our findings further support previous models in highlighting the relationship between tau and WMH but in addition, show temporal relationship and directionality that baseline tau is associated with future WMH burden.

The differences in our results between tau and WMHs from previous research could be attributed to limitations of the study such as sample size and statistical power, as our tau analyses included fewer participants compared to Aβ analyses, potentially providing greater sensitivity to detect Aβ WMH relationships. Heterogeneity in study populations may partly explain these findings. The present study included MCI and AD who may have higher tau burden compared to studies focusing on cognitively healthy participants, which could contribute to the detection of tau related effects on WMH. Additionally, our analyses were based on a two-year follow-up period, which may be insufficient to capture the full temporal differences of these pathological processes. Longer follow-up periods would provide greater insights into these relationships over time. Another important limitation in the present study was the limited number of participants with complete longitudinal data across all three measures (amyloid, tau, and WMH), which did not allow us to perform all analyses consistently in the same group of participants.

Findings from the present study highlight the relationships between WMH and amyloid and tau pathology, and the longitudinal analyses further demonstrate that these pathologies drive subsequent WMH accumulation rather than the reverse. The direction of these associations suggests that while WMHs may contribute to AD risk and potentially moderate disease progression, they are not the primary drivers of amyloid and tau pathology accumulation in AD. The regional specificity of the proteinopathy effects on WMH accumulation further supports the notion that posterior WMHs and their progression are likely linked to accumulation of AD rather than vascular pathologies.

## Data Availability

The data used in this study were obtained from the Alzheimers Disease Neuroimaging Initiative (ADNI) database and are available online at: adni.loni.usc.edu

https://adni.loni.usc.edu/

## Acknowledgments

Data collection and sharing for this project was funded by the Alzheimer’s Disease Neuroimaging Initiative (ADNI) (National Institutes of Health Grant U01 AG024904) and DOD ADNI (Department of Defense award number W81XWH-12-2-0012). ADNI is funded by the National Institute on Aging, the National Institute of Biomedical Imaging and Bioengineering, and through generous contributions from the following: AbbVie, Alzheimer’s Association; Alzheimer’s Drug Discovery Foundation; Araclon Biotech; BioClinica, Inc.; Biogen; Bristol-Myers Squibb Company; CereSpir, Inc.; Cogstate; Eisai Inc.; Elan Pharmaceuticals, Inc.; Eli Lilly and Company; EuroImmun; F. Hoffmann-La Roche Ltd and its affiliated company Genentech, Inc.; Fujirebio; GE Healthcare; IXICO Ltd.; Janssen Alzheimer Immunotherapy Research & Development, LLC.; Johnson & Johnson Pharmaceutical Research & Development LLC.; Lumosity; Lundbeck; Merck & Co., Inc.; Meso Scale Diagnostics, LLC.; NeuroRx Research; Neurotrack Technologies; Novartis Pharmaceuticals Corporation; Pfizer Inc.; Piramal Imaging; Servier; Takeda Pharmaceutical Company; and Transition Therapeutics. The Canadian Institutes of Health Research is providing funds to support ADNI clinical sites in Canada. Private sector contributions are facilitated by the Foundation for the National Institutes of Health (www.fnih.org). The grantee organization is the Northern California Institute for Research and Education, and the study is coordinated by the Alzheimer’s Therapeutic Research Institute at the University of Southern California. ADNI data are disseminated by the Laboratory for Neuro Imaging at the University of Southern California.

## Consent Statement

Written informed consent was obtained from participants or their study partner

